# The Seizure Embedding Map: A Spatio-Temporal Transformer for Comparing Patients by Ictal Intracranial EEG Features at Scale

**DOI:** 10.1101/2025.10.15.25338097

**Authors:** Akash R. Pattnaik, Zhongchuan Xu, William K. S. Ojemann, Carlos A. Aguila, Alfredo Lucas, Sarah Lavelle, Zack Goldblum, Peter D. Galer, Ryan Gallagher, Kathryn A. Davis, Nishant Sinha, Erin C. Conrad, Brian Litt

## Abstract

**Objective:** Planning invasive treatment for medication-resistant epilepsy relies on qualitatively interpreting seizure recordings from intracranial EEG (iEEG) recordings. Clinicians recommend treatment by mapping seizure onset patterns and locations, integrating multimodal data with their clinical experience and interpretation of the literature. Referencing a new patient’s seizures against past cases remains subjective, as implant strategies, electrode placement, and the electrodes’ relation to seizure onset vary across patients and centers. This study aims to rigorize this process by introducing a transformer model that embeds spatial and temporal information in iEEG recordings to categorize seizure networks and their relation to outcome across a large cohort of drug-resistant epilepsy patients. Our ultimate goal is to quantitatively compare multiple characteristics of new patients presenting for surgical intervention to thousands of prior patients to recommend best treatment.

**Methods:** We design and implement a custom spatiotemporal transformer that extracts features from iEEG seizure onset epochs. The model consists of convolutional layers that tokenize multi-channel iEEG, a spatiotemporal positional encoder that learns the relationship between sequences of tokens and the anatomical regions of the implantation to extract features across channels and time. Importantly, our model is flexible regarding to the number of iEEG contacts and the location of implants, being trained on both stereotactic EEG and electrocorticography implants. We validate seizure embeddings using unsupervised clustering to group seizure onset patterns using a cross-validated multi-class logistic regression model.

**Results:** The spatiotemporal model is applied to 882 clinical seizures from 102 subjects with drug-resistant epilepsy. Unsupervised clustering reveals 74 clusters of seizures that span multiple subjects, and a multi-class logistic regression model with 10-fold cross-validation reveals significant clustering of onset patterns in embedding space (validation accuracy = 0.8159). At the group level, seizures occurring closer in time exhibit more similar embeddings (*p* < 0.05), modeled with subject-specific random slopes and intercepts. Seizure clusters did not differentiate patients by therapy or postsurgical outcome, but they showed significant associations with the anatomical region of onset and seizure classification.

**Conclusions:** We propose a method for representing iEEG recordings of seizures with embeddings that contain spatial and temporal information. These embeddings can be characterized and compared across subjects to reveal common patterns in seizure onset. While this clustering did not separate patients by therapy and postsurgical outcome, there were significant associations with the anatomical region of onset and seizure classification. Future work will refine these methods to build a framework for characterizing seizures with deep learning incorporating multimodal data, including structural and functional imaging, semiology, patient history and demographics. We present this work as a first step toward quantitative, evidence-based decision making for patients with drug resistant epilepsy.

## Introduction

More than 51 million people suffer from epilepsy, a neurological disorder defined by multiple unpredictable and unprovoked seizures.^1^ Seizures occur when populations of neurons fire synchronously, in excess, and manifest as a multitude of semiologies ranging in severity from momentary lapses of awareness to convulsions, with an increased risk of injury and sudden unexpected death in epilepsy (SUDEP).^2^ Epilepsy care primarily aims to control seizures, though other co-morbidities are also managed as part of treatment.^3,4^ Anti-seizure medications (ASMs) are the first line of therapy, and 30-40% of patients fail two or more medications. For this population, neurosurgery or neurostimulation provide therapeutic alternatives, but both approaches require precise localization of the epileptogenic zone. To achieve this, clinicians use intracranial EEG (iEEG) to localize seizure onset and propagation regions. Current practice relies on qualitative interpretation of iEEG by multidisciplinary clinical teams. However, challenges in targeting the correct brain tissue and selecting appropriate surgical candidates limit long-term seizure freedom to 30-40% of these patients.^5^ Systematic and quantitative interpretation of ictal recordings offers the potential to improve localizations from iEEG and increase seizure freedom rates, yet methods for quantitative analysis remain limited.

Seizures recorded with intracranial EEG (iEEG) are traditionally interpreted through visual inspection by clinicians. Stereotyped onset patterns in frequency and amplitude can identify abnormal tissues. For example low-voltage fast activity (LVFA) is a common marker of seizure onset zone and a predictor of favorable surgical outcome.^6–9^ Other recognizable patterns include attenuation of background activity, rhythmic slow waves, and low-frequency periodic spikes, where occurrence can vary based on epilepsy subtype and distance to the seizure onset zone.^10,11^ Despite their clinical value, these patterns are typically detected through qualitative inspection by trained epileptologists, with low or poorly characterized intra-observer agreement.^12–14^

To reduce subjectivity, several quantitative methods have been proposed. The epileptogenicity index (EI)^15,16^ measures a ratio of high-frequency energy at seizure onset for each channel to identify brain regions that are likely epileptogenic. Dynamical system-based models can characterize seizure onset, propagation, and termination patterns based on their bifurcation types.^17^ Finally, network neuroscience-based models derived from iEEG data can uncover synchronizing and desynchronizing nodes in the epileptogenic network.^18,19^ While these techniques introduce objective biomarkers, they remain constrained by the spatial limitations of iEEG, and by assumptions about seizure models.

A major barrier to generalizability is the limited spatial sampling of iEEG. Patients are implanted with a combination of electrocorticography (ECoG) grids and strips placed on the cortical surface or in a growing number of US centers, stereotactic EEG (SEEG) depth electrodes that sample from deeper brain structures, white matter, and cortical tissue. The choice of implantation technique alters network-based measures of connectivity and their robustness.^20,21^ The quantity, density, and expansiveness of electrodes is associated with pre-implantation hypotheses of the focality of the epileptogenic zone.^22,23^ These variables make it difficult to integrate quantitative findings across patients, limiting our ability to identify shared seizure features or to relate seizure subtypes. Yet cross-patient comparisons offer the potential to reveal common ictal signatures, inform seizure classification frameworks, and to guide hypotheses for individualized therapies.

Deep learning offers a path forward and can be applied to overcome limitations in current state-of-the-art quantitative methods. As a sub-field of machine learning where weights and biases are tuned to learn patterns from training data, deep learning can leverage the wealth of data provided by iEEG recordings. Previous applications of deep learning with iEEG for epilepsy care have extended to seizure detection,^24–26^ interictal epileptiform spike detection,^27^ seizure prediction,^28^ and localizing the epileptogenic zone.^29^ Modern architectures such as transformers can uncover patterns in sequences of tokens over space and time, making them a compelling substrate for extracting salient features from seizure onset recordings in iEEG. Modern architectures such as transformers are particularly suited to iEEG, as they can model dependencies across space and time, extracting salient features from heterogeneous ictal recordings.

In this study, we develop and train a spatiotemporal transformer model for extracting features from ictal epochs of iEEG recordings. Our model contextualizes iEEG signal segments based on the anatomical region of interest from which they’re recorded, and the temporal order within the recording. The model can synthesize features from multiple seizure types and implantation strategies, enabling scalability to large numbers of patients and seizures from tertiary epilepsy centers. In this study, we apply a spatiotemporal transformer to a dataset of 882 seizures across 102 drug-resistant epilepsy patients to extract embeddings for each seizure. We hypothesize that similar seizure onset locations and patterns will cluster in the embedding space and that temporally clustered seizures will also have clustered features. This unsupervised approach enables data-driven pattern discovery and novel hypothesis generation, without bias from standard clinical annotations. Further, we posit that this approach can compare seizures across patients in a clinical setting, so that epilepsy care for new patients can draw from the collective experience with hundreds of prior patients.

## Materials and Methods

### Subject Information

Data from 102 subjects with drug-resistant epilepsy was retrospectively analyzed. Inclusion criteria for subjects included the following: 1) subjects received pre-implantation T1 MRI and post-implantation CT for localizing electrode contacts, 2) at least 1 clinical seizure was recorded with iEEG, and 3) clinical metadata on therapy, implantation type, targets, laterality, MRI lesion status, age, and sex was available. Following iEEG investigation, subjects received resection (n = 33) or ablation (n = 33) procedures and seizure outcomes after surgery were evaluated at 1 and 2 years using the Engel scale.^30^ All data collection was retrospective and approved by the Institutional Review Board of the Hospital of the University of Pennsylvania (HUP). Table 1 contains detailed demographic information on our study population.

**Table 1:** Subject Demographics.

| Category | Subcategory | n |
| --- | --- | --- |
| **Sex** | Female | 55 |
|  | Male | 47 |
| **Therapy** | Ablation | 33 |
|  | Neuromodulatory device | 22 |
|  | No surgery | 13 |
|  | Resection | 33 |
|  | Surg. planned | 1 |
| **Implant** | ECoG | 24 |
|  | SEEG | 78 |
| **Target** | Bilateral | 8 |
|  | Frontal | 14 |
|  | Insular | 4 |
|  | Multifocal | 8 |
|  | No surgery | 3 |
|  | Other | 8 |
|  | Temporal | 51 |
| **Laterality** | Unknown | 5 |
|  | Bi | 16 |
|  | L | 54 |
|  | R | 32 |
| **MRI Lesion** | Lesional | 53 |
|  | Non-Lesional | 49 |
| **Age at implant** | 10–19 | 5 |
|  | 20–29 | 33 |
|  | 30–39 | 24 |
|  | 40–49 | 27 |
|  | 50–59 | 12 |
|  | 60–69 | 1 |
| **Engel @ 12 M.** | 1 | 46 |
|  | 2 | 12 |
|  | 3 | 10 |
|  | 4 | 5 |
| **Engel @ 24 M.** | 1 | 40 |
|  | 2 | 7 |
|  | 3 | 10 |
|  | 4 | 7 |
| **Focal iEEG** | TRUE | 71 |
|  | FALSE | 31 |

### iEEG Data Collection

Intracranial EEG was conducted to localize epileptic networks. The intracranial electrodes (Ad Tech Medical Instruments, Racine, WI) included linear depth electrodes (1.1 mm in diameter with 5 mm spacing between contacts), as well as linear cortical strips and two-dimensional cortical grid arrays (2.3 mm in diameter with 10 mm spacing between contacts). The recordings were sampled at rates ranging from 256 to 1024 Hz, and signals were referenced to an electrode placed away from the suspected seizure sites, usually in the skull’s medullary cavity.

We used iEEG-recon, a validated and publicly available tool, to localize each electrode contact.^31^ Preimplantation T1-weighted MRI and post-implantation CT were linearly registered, and we transformed semi-automatically annotated electrode locations to MRI space. For each patient, we applied Freesurfer recon-all to register the Desikian-Killiany-Tourville (DKT) atlas to each patient’s native MRI space.^32^ To parsimoniously and grossly represent anatomical parcellations, we further aggregated regions of interest from the DKT atlas to consider 20 regions per hemisphere (mapping of the aggregated DKT atlas are provided in Supplementary materials).^22,33^To determine the mapping from each electrode contact coordinate to region, we applied a 5 mm radius around the center of each electrode coordinate and identified the region from the aggregated DKT atlas that most overlapped with the coordinate sphere.

Contacts that were exclusively in white matter were omitted from the study. Thus, each electrode contact from each recording was assigned a region of interest (Figure 1A).

**Figure 1.**
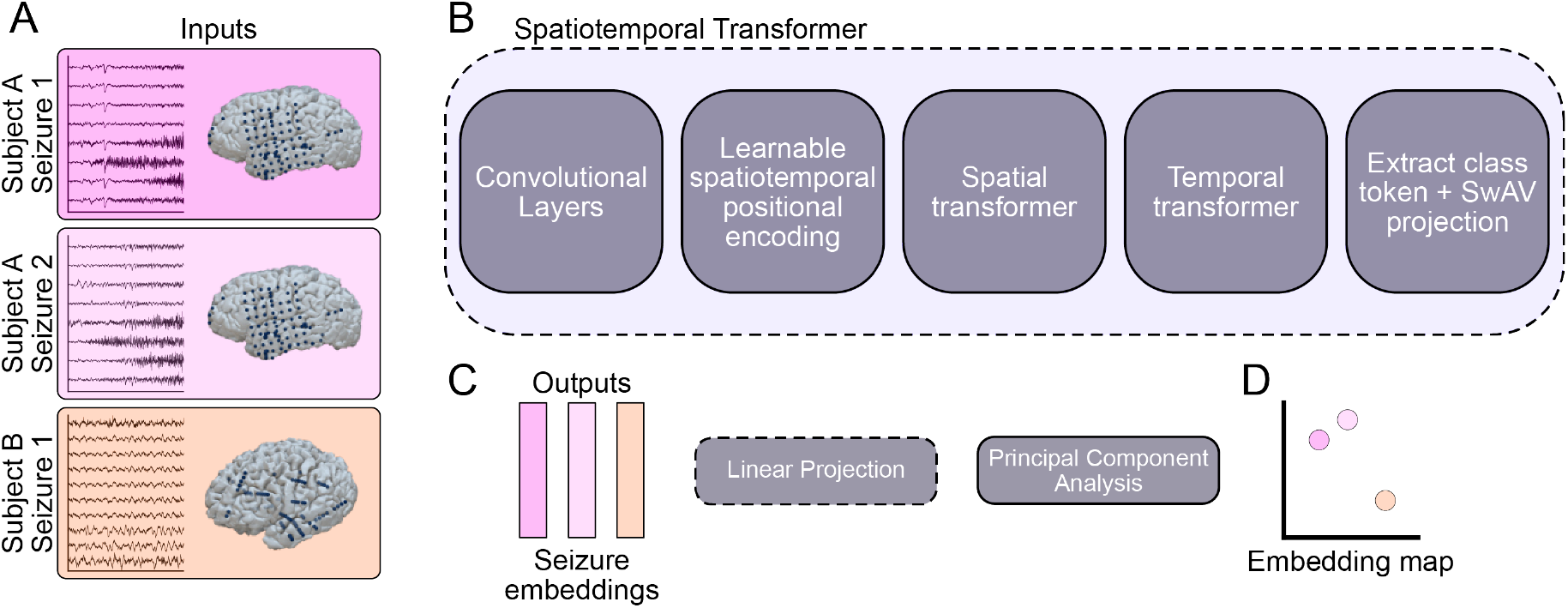
Methods overview. A) Input data consisted of iEEG recordings of seizure onset epochs and electrode locations, input as lists of DKT atlas regions where iEEG was sampled, derived from our iEEG Recon pipeline^31^. B) We designed and trained a spatiotemporal transformer model that consisted of convolutional layers for tokenization and time series feature extraction, learnable spatial and temporal positional encoding, sequential spatial and temporal transformers, and a learnable class token. We used SwAV (Swapping Assignments Between Views), a contrastive learning-based loss function, to train the model. C) The outputs consisted of 128-dimensional embedding vectors for each input sample. For task-specific analyses, we applied an additional linear projection head to fine-tune the embedding space for task-specific outputs, whereas in others we used the raw embeddings directly before PCA (Principal Component Analysis). D) To visualize model outputs, we plotted the embedding map: a scatter plot of each seizure sample in 2-dimensional, principal component space.

Seizure onset and offset were annotated by a board-certified epileptologist. In most cases, the earliest electrographic change was reported, though when this data was not available, we marked seizure onset as the unequivocal electrographic onset.^34^ Seizure onset epochs were selected as the first 10 seconds after onset recorded across all intracranial channels.

### Signal Preprocessing

Multi-channel iEEG recordings were preprocessed to isolate physiological signal and minimize noise. To each channel, we applied a second-order infinite impulse response filter with a quality factor of 30 at 60 Hz. The filter was applied forward and backwards to avoid artificially introducing a phase offset in the signal. Data were bandpass filtered between 0.5 Hz and 120 Hz with a 10th order Butterworth filter to remove DC offset and slow drift as well as high frequency noise near the Nyquist frequency, to reduce aliasing. We re-montaged data to a bipolar montage, where signals from adjacent contacts were subtracted. Contacts were subtracted within SEEG leads and ECOG grids to maintain comparable local distance between bipolar pairs. To ensure input to the spatiotemporal transformer model was uniform, we downsampled signals to 256 Hz from their original recording frequency using polyphase phase filtering and we padded the number of channels up to 150 total channels with measurements of 0 Volts at each time point (Figure 1A).

### Model Architecture

EEG signals are continuous by nature. Thus, the first component of the model consisted of cascading 1-dimensional convolutional layers to tokenize input signals and encode features. Convolutional layers were applied independently to each channel of each multi-channel iEEG recording. The convolutional feature encoder followed the CNN architecture from *wav2vec 2*.*0*^35^, with number of channels (64, 128, 64, 32), strides (5, 2, 2, 2), and kernel widths (10, 3, 3, 2). An adaptive pooling layer followed the convolutional layers to ensure that the output shape was the same size across varying input lengths to reduce the input to 20 tokens.

After the input EEG samples were tokenized by the convolutional layers, we applied spatiotemporal positional encoding. Positional encoding in the standard transformer architecture have been used in NLP (Natural Language Processing) to provide context to the sequence of tokens to ensure that the relationship between adjacent words are maintained in parsing sentences. We applied this same strategy to iEEG recordings to preserve both the temporal structure of the signals in the order of tokens and the spatial relationships between recording channels. Thus, we encoded each token position in the sequence and the region index of the aggregated DKT atlas region from which each channel was recorded. We used learnable positional encoding, allowing the model to infer positional relationships directly from data rather than imposing predefined temporal structures (as sinusoidal positional encoding). Positional encoding information was added to the output of convolutional layers and learned over training.

To aggregate information across both spatial and temporal dimensions, we implemented a sequential spatial-temporal Transformer encoders. The first spatial Transformer computed attention across channels to capture spatial dependencies among feature maps. Each channel output from the CNN encoder was treated as a token. A two-block Transformer encoder was applied with an internal embedding size of 640, 7 attention heads, 64 inner-head dimensions, and a 2048-dimensional feed-forward layer. An attention mask was used to exclude padded channels from training. The output of the spatial Transformer was then rearranged so that each token corresponded to a temporal segment (a sampled window in time), represented by the aggregated features across all channels at that time point. The second temporal Transformer encoder was applied over these temporal tokens to model temporal dependencies, consisting of 4 Transformer blocks, an internal size of 4800, 8 attention heads, 64 inner-head dimensions, and a 2048-dimensional feed-forward layer.

We implemented a learnable class token to represent the embedding of each sample. The class token is a token appended to the input to the temporal transformer, and we extract this token after learning. The parameters in the class token vector were learned during training and extracted to condense each sample’s transformer output. The class token was passed to a projection head, which normalized each vector and applied a linear layer for the loss function. This ensured the output representation for each sample was a unit vector with size 128 (Figure 1B).

After self-supervised pretraining, we froze the encoder weights and trained a new linear projection head, a lightweight classification layer that maps the pretrained representations to task-specific outputs, using patient class labels to perform task-specific classification and evaluation. This fine-tuning step adapts the pretrained embeddings to the downstream classification task while preserving their intrinsic structure learned during self-supervised training.

### Model Training

As the objective of our model was to learn unsupervised representations of seizure activity, we trained it using a contrastive self-supervised learning approach. Contrastive learning teaches the model to bring similar examples closer together in the embedding space while pushing dissimilar examples apart. For each 10-second seizure-onset recording, we generated multiple augmented views of the same signal to create training pairs. Specifically, we randomly extracted six different segments (“views”) from each recording across all channels, two longer 7.81-second segments (2,000 samples at 256 Hz) and four shorter 3.91-second segments (1,000 samples at 256 Hz). These longer and shorter views encouraged the model to capture both coarse and fine temporal structure in the iEEG data. Each “view” was drawn from a random temporal offset within the original 10-second window. During training, “views” originating from the same seizure served as positive pairs (expected to have similar representations), while segments from different seizures or patients served as negative pairs. This design encouraged the model to learn seizure-specific features that generalize across time and patients. We optimized the network using the Swapping Assignments Between Views (SwAV) loss function.^36^ SwAV extends contrastive learning by maintaining a queue of previously seen samples, that acts as an extended pool of negative examples beyond the current batch. This improves representation diversity without requiring very large batch sizes. The queue was activated after 50 epochs with a pool size of 256 samples. The model was trained for 500 epochs with a batch size of 32 using the Adam optimizer with learning rate of 0.01. All experiments were performed on an NVIDIA A40 GPU (CUDA 12.1, 48 GB VRAM).

### Statistical Analysis

We estimated the variability in training performance by training the spatiotemporal transformer over 500 epochs and varying the random seed over 10 iterations. We subsampled the number of channels to determine the spatial resolution at which clustering can be preserved and re-trained the model with 5 seeds per subsampling resolution. To determine if the clustering of clinical variables was significant, we permuted labels and computed the mean distance from the centroid of each clinical cluster. We rejected the null hypothesis if the mean distance from the centroid was lower than the 95th percentile mean distance over 1000 bootstrap iterations.

Each seizure embedding vector, the output of the model, consisted of 128 learned features per 10-second seizure-onset event (Figure 1C). After training, we applied a linear projection layer with an output shape of 128, fine-tuned using seizure-onset zone labels to adapt the embedding space for task-specific visualization. This information was further condensed using Principal Component Analysis and visualized in two dimensions with a scatter plot, where each point represented a 10-second seizure-onset event (Figure 1D). We tested the association between the temporal distance of two seizures from the same patient and their embedding distance using a random-slope, random-effects model, with time difference as the regressor, embedding distance as the predictor, and patient as the random effect. Each sample for this analysis was a pair of seizures, and we visualized the fixed and random effect slopes and intercepts with line segments.

## Data Availability

Seizure epoch data, saved in EDF file format, and model weights are available at Pennsieve.io.^37^ The model architecture and other analysis code is available on Github. All of the above resources are available here. A visualization and iterative tool of the embedding generated in this study is available here.

## Results

### Training the Spatiotemporal Transformer with Ictal iEEG

We constructed a spatiotemporal transformer model with convolutional layers for time series feature extraction, learnable positional encoding of anatomical regions and token sequence, and a learnable class token that represented the embedding of each sample. The model was trained over 500 epochs and applied to 882 seizures from 102 subjects with drug-resistant epilepsy. SwAV loss was minimized to train the model, and we tracked the loss over 10 seeds that randomized initial model weights. Across seeds, we observed consistent decrease in SwAV loss, with a decrease in loss at 50 epochs when the queue of negative examples for SwAV to evaluate became active (Figure 2A). We extracted seizure embeddings from the learnable class token after 500 training epochs. To determine how similar seizure embeddings were across model initializations, we first computed pairwise cosine similarity for the seizure embeddings at each seed. Second, we computed the Pearson correlation between cosine similarity matrix to answer how consistently seizures were evaluated as similar or dissimilar across initializations (Figure 2B). We observed consistent and moderate correlations between seizure embeddings across initializations apart from one outlier, seed 5. Thus, we concluded that our model consistently embedded seizures across training initializations.

**Figure 2.**
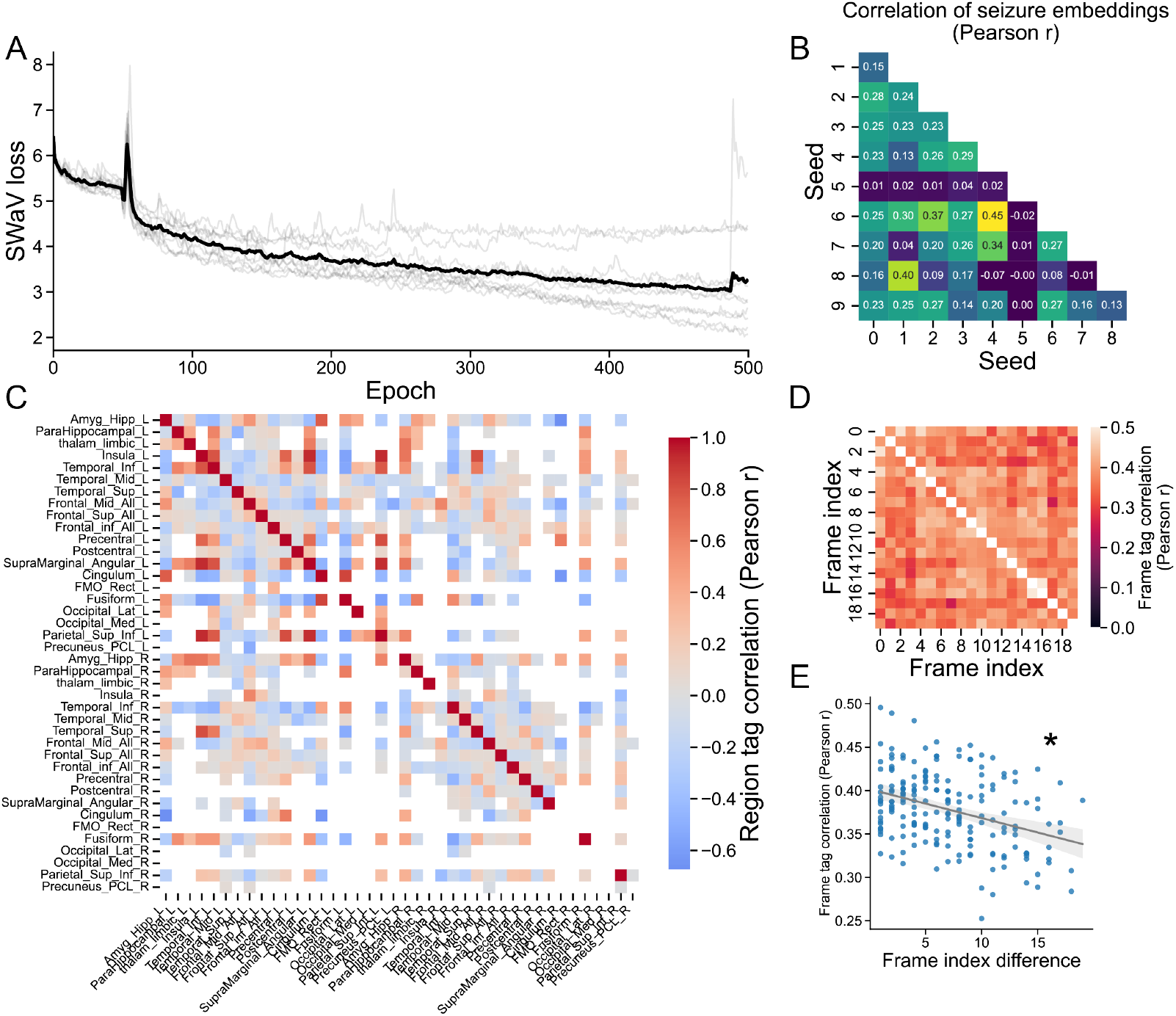
Model training and evaluation. A) We trained the spatiotemporal transformer over 500 training epochs and tracked SwAV loss at each epoch. Ten random weight initializations (seeds) were applied, and loss is shown in light gray lines. The mean loss across the ten initializations is shown in black. We observed consistent and moderate correlations between seizure embeddings across initializations. B) For each seed, we computed the cosine similarity between seizure embeddings at the final training epoch. The Pearson correlation between seizure embedding similarities indicates the consistency of embeddings at the end of training, where most seeds had similar seizure embeddings with seed 5 as an outlier. C) We computed the Pearson correlation between positional encoding tags for each anatomical region. Pearson correlations that fluctuated around 0 were omitted as they were likely noise. For all other pairs, red indicates more positive tag correlations and blue indicates more negative tag correlations. D) The model learned a tag for each frame (temporal sequence of tokens), and we computed pairwise Pearson correlation between the frame tags. E) Frames that were closer together in sequence had more similar frame tags (*r* = −0.36, *p* < 0.05). This suggests that temporally adjacent seizures have more similar temporal assignment.

The spatiotemporal positional encoder assigned a tag to each frame (EEG segment within the temporal sequence of the entire input clip) and anatomical region (mapping of each contact to a custom brain imaging atlas). We sought to characterize how the model learned the encoding of tokens using the correlation between tags, similar to approaches that have been used for learnable positional encoding with vision transformer models.^38^ Pearson correlations between region tags show similar representations of left mesial temporal structures and occipital/parietal regions (Figure 2C). We applied a similar analysis to the tags for each frame, where frame 0 is the first segment for each sample and frame 19 is the last time (Figure 2D). We found that frames that are closer together in time have similar tags (Figure 2E; *r* = −0.36, *p* < 0.05), suggesting that adjacent EEG segments have similar temporal assignment. To simulate missing electrodes, we conducted an ablation study by randomly removing channels and computing the cosine similarity between the full and masked channel embeddings. The resulting similarity scores are reported in the Supplementary Material (Figure 9). We found the model’s embeddings are highly robust to small channel loss (≤ 10%), but representation quality degrades rapidly beyond 30% masking, indicating that spatial completeness of EEG input remains crucial for consistent seizure representation.

### The Seizure Embedding Map Reveals Clusters of Seizures Across Subjects

To characterize how seizures cluster across patients, we applied an unsupervised clustering algorithm on the seizure embeddings. We applied k-means clustering over a range from k = 2 to k = 256 and computed the Silhouette score at each interval. We determined that the optimal number of clusters was 74 by finding the first interval where the Silhouette score passed 0.5 (Figure 3B). Projecting the embeddings to 2-dimensional space using principal component analysis (PCA) and mapping each seizure to its cluster qualitatively revealed cluster quality and “zones” of the seizure embedding map (Figure 3A). We found that clusters were comprised of seizures across subjects (median number of subjects per cluster = 2, IQR = 2, Figure 3C) and multiple seizures (median number of seizures per cluster = 6.5, IQR = 10.75, Figure 3D). Thus, applying unsupervised clustering to the spatio-temporal transformer-based embeddings revealed clusters that spanned subjects and their customized spatial sampling of iEEG contacts. The embedding space visualizations after each architectural block are provided in the Supplementary Material (Figure 8).

**Figure 3.**
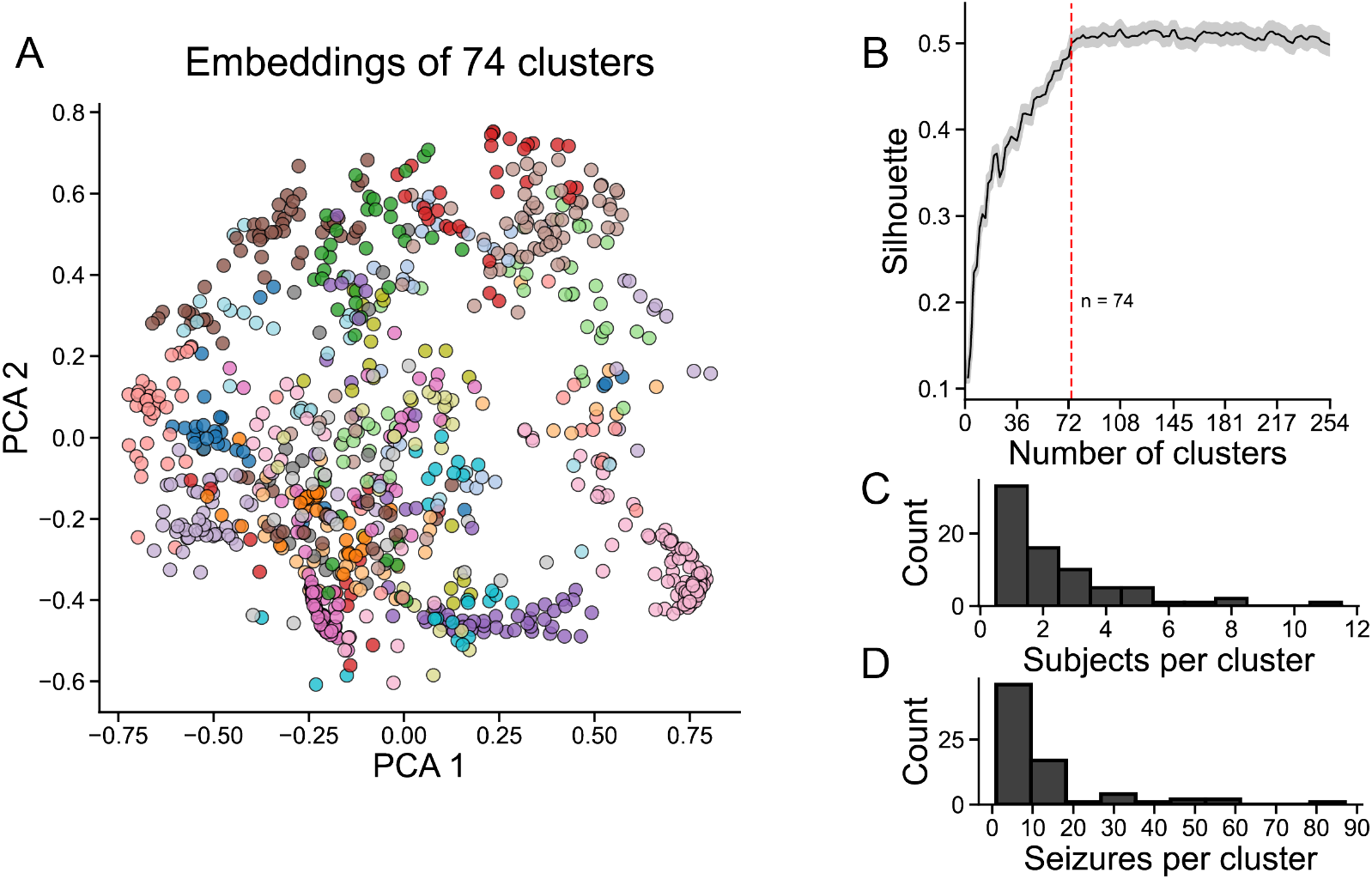
Applying unsupervised clustering on seizure embeddings. A) We transformed the 128-dimension embeddings into 2-dimensions using principal component analysis. The first and second principal component coefficients are shown. Points represent each seizure embedding (n = 878) and are colored based on cluster assignments for 74 clusters using k-means clustering. B) 74 clusters (vertical red dashed line) were determined by computing the minimum number of clusters that is above a silhouette score of 0.5. The mean and standard deviation across samples for each number of clusters is shown in the black line and grey envelope. C) The histogram shows the number of subjects represented by each cluster with number of subjects per cluster on the x axis and the number of clusters on the y axis. D) The histogram shows the number of seizures represented by each cluster with number of seizures per cluster on the x axis and the number of clusters on the y axis.

### The Seizure Embedding Map Characterizes Seizure Similarity Within Subjects

Previous studies have aimed to compare seizures within subjects, to uncover patterns in seizure types and determine typical and atypical seizures which can be used for epilepsy surgery planning. For each subject, we computed the average pairwise cosine similarity for all pairs of seizures recorded from that subject (Figure 4A-B). We highlight two example subjects to illustrate variability in seizure similarity. Subject 1 had the lowest average pairwise similarity (avg. similarity = 0.11), with two recorded seizures that spanned two different clusters. Qualitative review of the seizure recordings reveals different onset patterns and timings in the two seizures. The first seizure (Figure 4C) consists of lower amplitude and later onset relative to the second seizure (Figure 4D) which also features pre-seizure spiking. In a second subject (Figures 4E-G), we found similar pre-seizure chirps and onset channels across three seizures.

**Figure 4.**
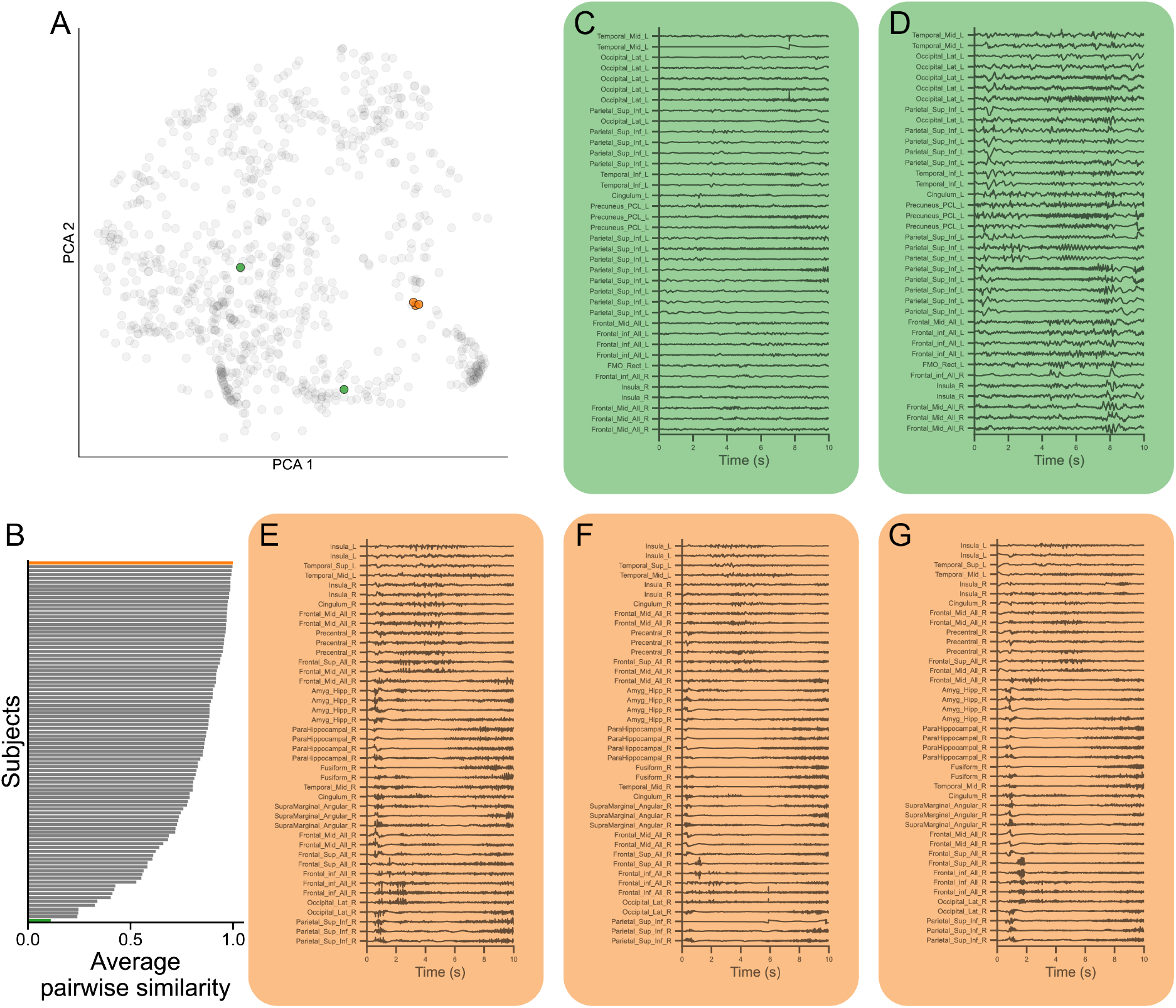
Comparing seizures within subjects. A) We highlight two example patients with low (green) and high (orange) pairwise seizure similarities in PCA space. B) For each subject, we computed the pairwise seizure similarity. C) The subject with low pairwise seizure similarity had two clinical seizures, we visualize the EEG with channel labels as the anatomical region information that was input into the model for the positional encoder. D) We repeat the EEG visualization for the subject with high pairwise seizure similarity.

### Seizures with Similar Onset Locations are Captured by the Seizure Embedding Model

To further investigate how onset locations are represented on the seizure embedding map, we projected clinically annotated seizure onset regions onto the low-dimensional seizure embeddings (Figure 5A-D). We found that left frontal (n = 413), left temporal (n = 245), right frontal (n = 19), and right temporal (n = 109) onsets were all significantly clustered by bootstrapping the mean distance to the centroid over 1000 iterations (*p*_*Bonferroni*_ < 0.05 for all four onset locations). A 10-fold cross-validated linear classifier to perform four-class classification was moderately accurate (acc. = 0.816) at predicting the onset location (Figure 5E). An ablation study using null models was conducted on the same set of seizures to evaluate the effectiveness of the spatial encoding. Specifically, one null model employed only the seizure embeddings (with the spatial encoding removed), while the other used only the spatial encoding (without the seizure data), yielding classification accuracies of acc. = 0.456 and acc. = 0.528, respectively.

**Figure 5.**
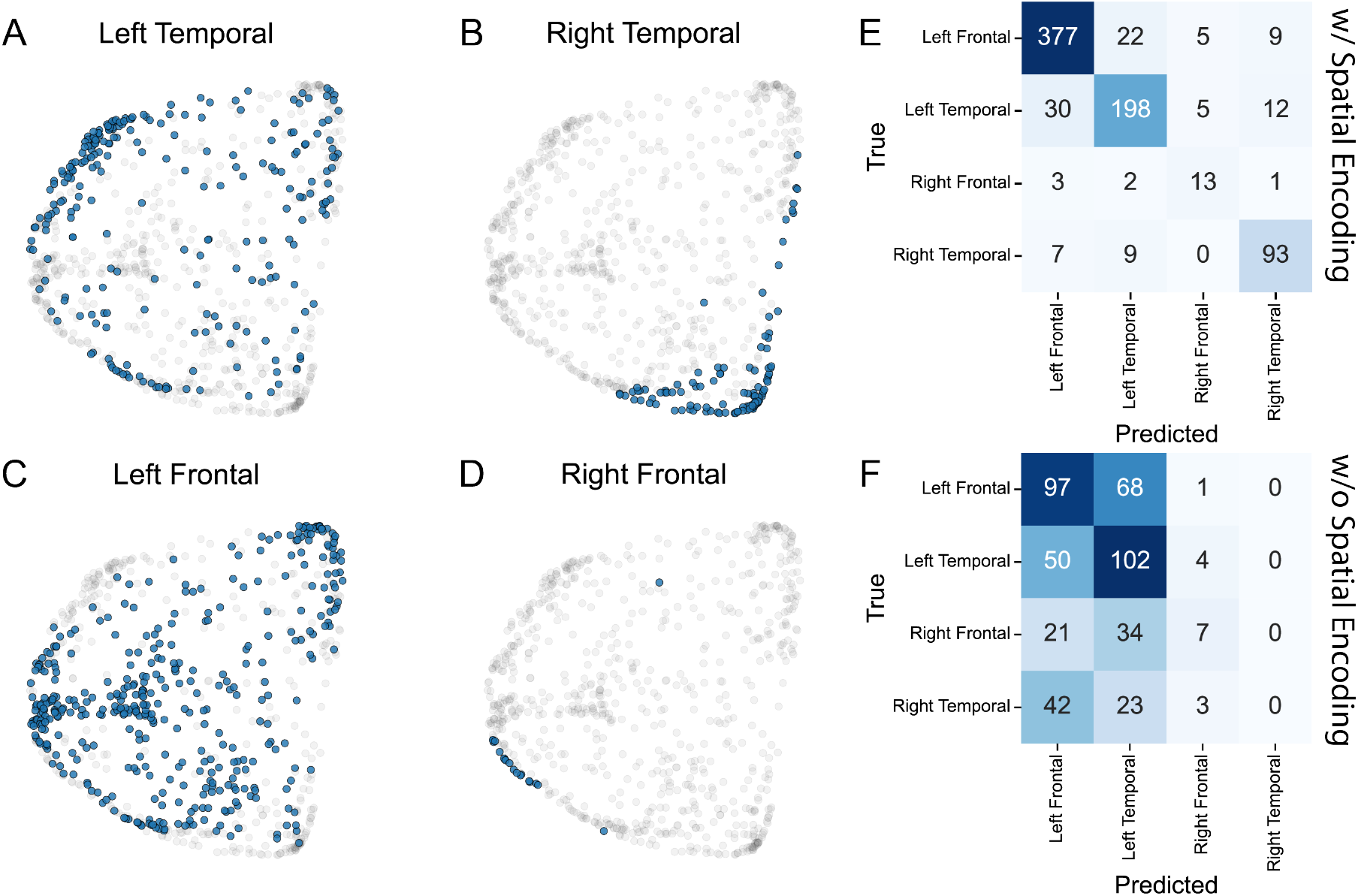
Projecting seizure onset locations onto embedding map. A) We projected labels of the anatomical region of seizure onset onto seizures and highlighted Left Temporal onset seizures in blue against all seizures in grey. Each dot is a seizure in 2-dimensional PCA space. B-D) We repeated the same methodology as A for Right Temporal, Left Frontal, and Right Frontal onset seizures. E) Confusion matrix for linear evaluation with 10-fold cross validation to predict seizure onset location using seizure embeddings as features (acc. = 0.816). F) A null model using the same architecture and methodology, but with spatial encoding removed (acc. = 0.456).

### Temporally Clustered Seizures are Characterized as Similar Embeddings

Seizure clusters are known to exhibit similar network dynamics as they progress from onset, propagation, and termination phases.^39^ Previous literature has revealed positive correlation between temporal offset in seizure timing and seizure dissimilarity.^40,41^ Thus, we asked if our derived seizure embeddings were sensitive to seizure clusters. For each pair of seizures recorded from the same subject, we computed the temporal offset and the cosine-similarity based distance between seizures. We applied a random-slope random-intercept model with the subject as the random effect to model cohort-level associations while accounting for subject-level trend differences. The fixed effects revealed a significant positive association between embedding distance and time difference (*p* < 0.05, Figure 6A). To further understand how each subject’s seizure pairs were represented in the model, we visualized the seizure pairs and slope and intercept of the fitted model for that subject for the two subjects with the highest (Figures 6B-C) and lowest associations (Figures 6D-E).

**Figure 6.**
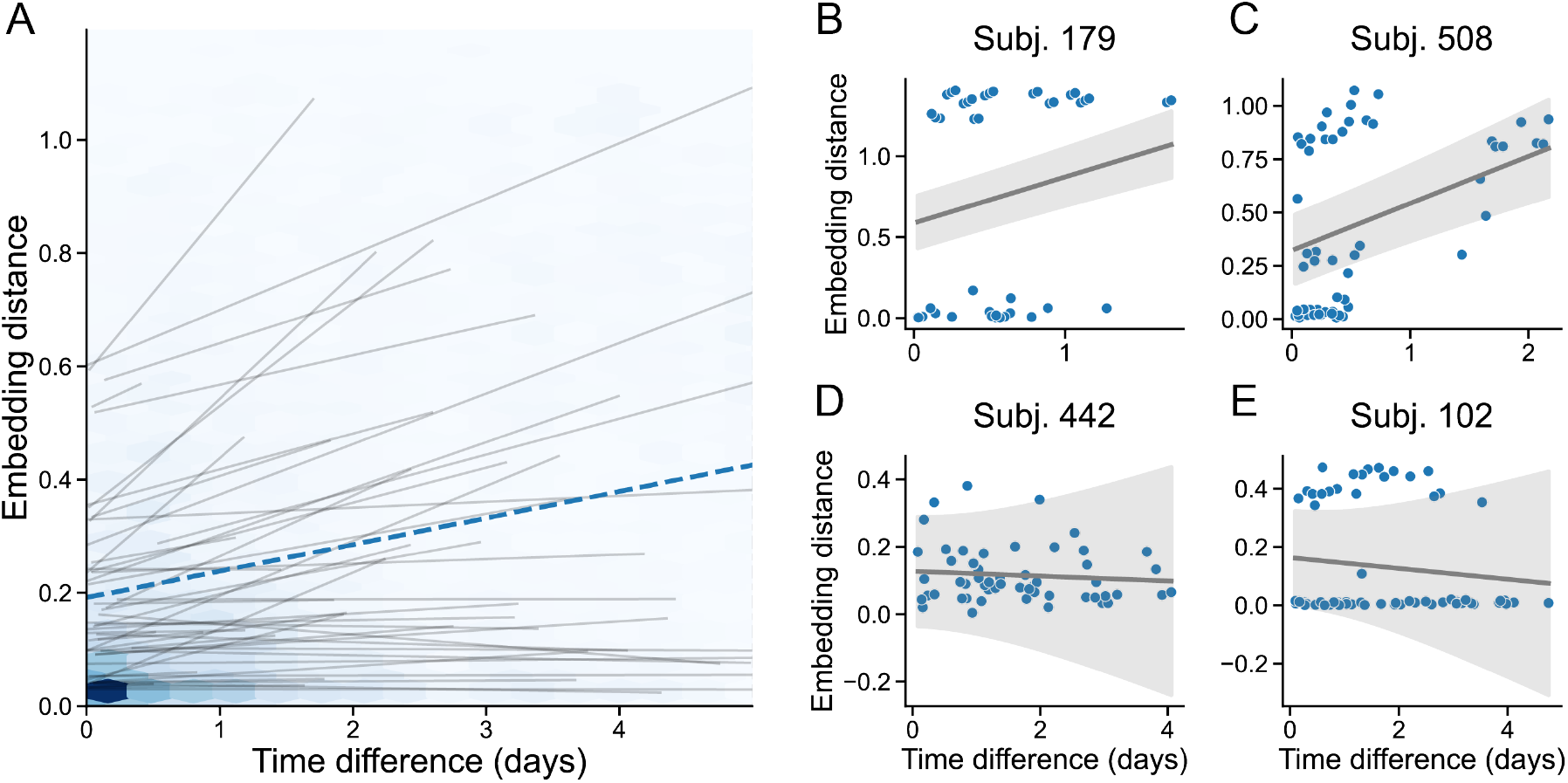
Associating temporal difference in seizures with embedding distance. A) To model trends between patients, we fit a random effects model with time difference as the regressor and embedding distance as the outcome with subject ID as the random effect. We visualize each data point using a hexbin plot, where darker blue hexagons indicate more samples in that region. The dashed blue line shows the fixed effect slope and intercept, and the thin grey lines show each subject’s random effect. B and C) Scatterplots show a given subject’s seizure pairs and the subject’s random slope and intercept with standard error of the mean in the envelope derived from the variance in the slope and intercept. B and C show the subjects with the most positive slope. D and E) Similar scatterplots as B and C but for the subjects with the most negative slope.

### Seizure Embeddings are Not Sufficient for Clustering Subjects by Therapy and Outcome

We finally tested if similar patients had similar therapies and outcomes after treatment. To summarize a subject’s representation, we computed the vector sum and maxpool over all of that subject’s seizure samples and normalized the sum by the norm to ensure that all subject embeddings were unit vectors. We then determined if each subject’s embeddings were clustered relative to two clinical variables: chosen therapy, and seizure freedom after resection and ablation procedures. For the therapy analysis, we limited our analyses to subjects who received resection, ablation, and neuromodulatory device, omitting those who did not receive surgery after iEEG explantation. For the seizure freedom analysis, we limited our analyses to subjects who received resection or ablation procedures. We iterated the number of clusters parameter to capture multiple resolutions of clustering and computed the similarity of data-driven hierarchical clusters of subject embeddings with the clinical labels. We observed clustering similarity, measured by the adjusted Rand index, across scales for both therapy and seizure freedom (Supplementary Material Figure 7A) and visualize the contingency table between hierarchical cluster assignments and seizure freedom for the best clustering resolution (Supplementary Material Figure 7B). Thus, embeddings derived from seizure onset recordings are not sufficient for assigning patients to clusters based on clinical variables of interest, and the addition of more, multi-modal features may be required to better capture heterogeneity in cohorts based on choice of therapy and seizure freedom.

**Figure 7.**
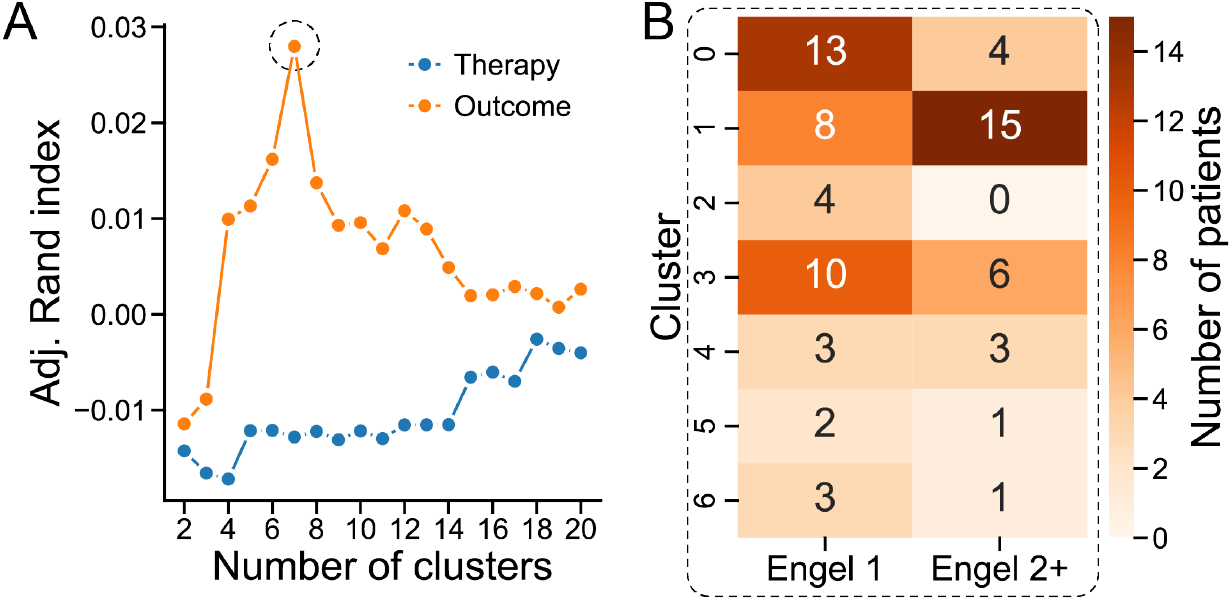
Evaluation of clustering resolution and alignment with seizure outcome. A) Clustering similarity across different resolutions, measured by the adjusted Rand index, for therapy and seizure freedom. B) Heatmap of the contingency table comparing hierarchical cluster assignments with seizure freedom (Engel 1 and Engel 2+) at the optimal clustering resolution.

## Discussion

The goals of this study were to develop a novel, objective, quantitative method to compare intracranial EEG recordings from a new patient to the common experience with many patients, leveraging state of the art deep learning and computer vision techniques. A second, more clinical goal was to determine if this method could predict which therapy was likely to be most effective for an implanted patient, based upon 10 second seizure onset epochs from the iEEG alone. Our findings indicate that the method has promise for clustering patients and seizures, and for comparing new patients to the common experience. Similar onset localizations across patients were more likely to be clustered together, as were seizures that occurred closer together in time for individual patients. These groupings alone were not deemed significant enough to be used alone clinically, but they validate common clinical findings used in clinical practice during manual review. The finding that 10 second seizure onset segments are not sufficient to predict outcome from therapy is not surprising, given the complexity and heterogeneity of epilepsy subtypes, sampling error in intracranial EEG and the other factors that go into clinical decision making about therapy, including history, genetics, brain imaging, seizure semiology, neuropsychological testing and demographics. Still, the method validates several clinical findings, and is expandable to include these disparate modalities, which we believe will, together, have much stronger clinical impact. In addition, adding much more data from scalp and iEEG recordings, during, leading up to, between and after seizures, has been shown to increase predictive value for clinical use, and we expect that also to be true for our new method.^42^ For these reasons we find our results above to be encouraging but there are also technical considerations worthy of discussion, particularly given the nonuniformity of data used in tertiary epilepsy care.

Embedding seizure recordings from intracranial EEG poses several challenges: implantation strategy varies between subjects, seizure onset patterns differ across patients and epilepsy types, and validation of embedding methods are poorly established. In this study, we use transformer models to overcome these challenges and present several novel contributions to the domain of characterizing seizure epochs from intracranial EEG. We develop a transformer model that operates on EEG data without necessitating manual feature extraction. We apply spatial and temporal positional encoding to contextualize EEG from where and when it is recorded. The spatial encoder leverages known differences in EEG properties across anatomical regions.^33,43,44^ We demonstrate seizure embeddings with similar onset regions are similar, and seizure embeddings that are temporally proximal are similar, two known clinical findings that validate the method. We also demonstrate the robustness of our method with several analyses, including an “ablation” study, showing its sensitivity to the number of electrodes used, and a robustness analysis. We believe this novel approach will stimulate further innovation in autonomous methods for comparing seizures, including those that may be expandable to more data modalities and other medical challenges for classifying patients, predicting outcome and guiding therapy. More immediately it is our hope that the method may stimulate new approaches that can be used in implantable or wearable devices, both in and outside of hospital.

### Comparing Our Model with Other EEG-Based Deep Learning Approaches

Applications of deep learning to EEG data and epilepsy span supervised and unsupervised learning tasks applied to epileptic network localization, seizure detection, and seizure warning. The cost of obtaining large, high-quality iEEG seizure datasets recorded across heterogeneous conditions of epilepsy types, seizure types, and implantation have limited supervised learning methods, which rely on such labels to learn and generalize effectively. In this study, we used a self-supervised and contrastive learning-based loss function to characterize seizures. SwAV learns by comparing positive sample pairs (EEG augmented views from within the same seizure) and negative sample pairs (EEG augmented views from across seizures and patients), thereby transforming an unsupervised task into a supervised one. Desai et al. pursued a similar approach with contrastive learning to identify similar recordings from the responsive neurostimulator device.^45^ Their approach consisted of a preceding unsupervised clustering step to identify positive pairs (within cluster samples) and negative pairs (cross-cluster samples). By contrast, our methodology assumes that the underlying distributions of signals are preserved over time by taking multiple temporal crops of a single sample—recordings from the same location at different times should be more similar than recordings from different locations and patients. SwAV also condenses and discretizes each class token into a prototype, an N-dimensional unit vector that’s similar to a pre-determined basis vector. Other groups have also relied on unsupervised learning for seizure detection and spanned applications of deep learning and non-deep learning models.^25,26,46–48^ Adopting unsupervised learning and achieving comparable performance to supervised learning holds promise for seizure annotation and detection without necessitating large annotated datasets.^49,50^ Further research should focus on characterizing loss functions for EEG and improving interpretability of self-supervised models.

### Seizure Clusters, Onset, and Evolution

We hypothesized that seizures that cluster temporally would have more similar embeddings. Previous work has found that seizure pathways, non-linear evolutions of functional connectivity over the course of a seizure, vary on linear and circadian temporal scales.^40,41^ Indeed, we found that pairs of seizures that had short inter-seizure intervals had similar embeddings. Our findings align with evidence from animal models, where seizure clusters share similar properties, such as hippocampal spike rate.^51^ Seizure clusters also exhibit trends of shorter duration and heighted severity as clusters progress, suggesting that combining deep-learning based seizure embeddings with clinical scales of severity may be informative.^52,53^ Other studies have developed biomarkers that are sensitive to focal only and focal-to-bilateral-tonic-clonic seizures, and the study of how embeddings encode features of severity should be investigated.^18^ Our results revealed that seizures with similar onset locations cluster together. Sensitivity to seizure onset zone is an important factor to consider when developing a seizure embedding model^54^—surgical planning relies on seizure onset channels to identify foci in the epileptogenic network. This finding also suggests that building models that are aware of spatial sampling can improve clinical relevance. Overcoming sampling bias remains a challenge for iEEG research,^21,55,56^ and our approach aims to account for sampling bias through positional encoding. Certainly, other approaches, such as graph neural networks^29,57,58^ and integration with other data modality such as imagings, can add spatial context into model inputs and should be tested with seizure recordings.

### Clinical Implications of Cohort-Level Seizure Embeddings

Embedding vectors that automatically extract features from high-dimensional iEEG present the opportunity to change epilepsy care. Prior literature has aimed to develop features that can distinguish between good and poor candidates for surgery and device implantation, based on seizure outcome and focal epilepsy networks.^22,33, 59–63^A robust, multi-patient, and multi-center model could potentially automatically uncover these features while ingesting large amounts of raw data.^64,65^ Our model shows initial promise, though our inability to significantly cluster subjects based on therapy choice and outcome could be due to the small cohort size of patients, all originating from a single center. It may also be due to the limited and single modality data entered into the model, as discussed above. Future work could apply such models or theories to larger, multi-center and multi-modality datasets to integrate clinical knowledge across epilepsy centers. Perhaps similar patients at certain centers are more responsive to therapy than those from other centers. Uncovering such trends could improve clinical practice across epilepsy centers globally, just as stereotactic EEG implantation gained popularity in North America after being developed in Paris by Jean Talairach and Jean Bancaud during the 1960s.^66^ Rapid and informed adoption of new clinical innovations in epilepsy care could ultimately improve the quality, rigor and uniformity of therapy for patients with drug-resistant epilepsy.

### Limitations and Future Work

Our work has important limitations at this early stage. Quantitative methods to extract features of epileptogenicity from intracranial EEG seizures are limited by clinical and spatial constraints. The choice to implant iEEG is exclusively guided by clinical need, patients who have well localized epilepsies from non-invasive imaging may not be indicated for iEEG, causing an inherent bias in a dataset of iEEG seizures. We limited input data to the first 10 seconds of a seizure, and varying the window of data slightly could influence the temporal encoding of input data. The potential variability in markings of the unequivocal electrographic seizure onset with respect to the start of the recording could vary, as the earliest electrographic change period can last a variable amount of time.^34^ We did not include data beyond the first 10 seconds of seizures, and additional data could potentially be beneficial for comparing seizure recordings.

Additionally, improved encoding methods and constraints could further enhance embedding generation. For instance, using a graph neural network could compensate for missing observations, while incorporating causal representation learning techniques would ensure the embedding space is disentangled, which will help our capability to address the challenges of sampling bias and cross-patient differences. This model was developed for experimental purposes on data from one epilepsy center and has not been cross validated in a multi-center dataset.

Future work towards clinical translation should amass data from multiple data modalities, from multiple epilepsy centers globally, to overcome site specific biases in patient selection, spatial sampling, and epilepsy management. This will require extensive cross-validation of the deep learning model as well as validation against known variability in seizure onset dynamics. The field of deep learning is evolving rapidly, as novel techniques are discovered, and computational resources become more commonplace. Future work should also consist of developing pipelines to test and deploy deep learning models using cloud-computing, to ensure that state-of-the-art quantitative methods are accessible. Another major conclusion is the need for centralized or linked data repositories spanning hundreds to thousands of patients, which will greatly enhance the ability of deep learning methods to contribute to clinical care. Efforts to do this are underway, such as Epilepsy.Science, Pennsieve,^37^ Open Neuro^67^ and BDSP (Brain Data Science Platform).^68^ Developing routine systems where data from major centers is automatically shared as part of routine clinical practice could dramatically accelerate these efforts and dramatically improve patient care.

## Conclusion

We present a spatiotemporal transformer model for embedding and comparing iEEG seizure recordings across a large cohort of drug-resistant epilepsy patients. Our findings show that seizure embeddings capture clinically meaningful features, clustering by anatomical region of onset and seizure classification. By enabling quantitative comparisons of seizures across patients, this approach lays the groundwork for data-driven discovery of biomarkers, novel phenotypes, and developing more rigorous frameworks for surgical decision-making and ultimately more general patient care. Future work should build upon these findings by integrating multimodal data, such as imaging, semiology, and patient history, to improve generalizability and advance toward evidence-based treatment recommendations informed by thousands of prior cases.

## Data Availability

All data produced in the present work are contained in the manuscript

https://github.com/sirrus98/ieeg_sz_embedding

## Funding

B.L recieved support from the National Institutes of Health (NIH) (NINDS R01 NS125137 and NINDS DP1 NS122038) and Mirowski Family Foundation; by contributions from Neil and Barbara Smit; and by contributions from Jonathan and Bonnie Rothberg. E.C.C received support from the National Institutes of Health National Institute of Neurological Disorders and Stroke (NINDS K23 NS121401-01A1) and the Burroughs Wellcome Fund. N.S. received support from the National Institutes of Health National Institute of Neurological Disorders and Stroke (NINDS K99 NS138680). K.A.D received support from the National Institutes of Health National Institute of Neurological Disorders and Stroke (NINDS R01 NS134625-01). W.K.S.O. was supported by the National Science Foundation Research Grant Fellowship (DGE-1845298). C.A.A. received support from the National Science Foundation Research Grant Fellowship (DGE-2236662).

## Ethical Statments

This research study complies with Declaration of Helsinki. iEEG data recordings were collected as part of routine clinical care from 102 patients at the Hospital of the University of Pennsylvania (HUP). The collection of iEEG data for research purposes was approved by the HUP Institutional Review Board. Informed consent was obtained from each subject. The approval number for this study is 811097, titled ‘iEEG’.

## Competing Interests

The authors have no potential conflict of interest with the present study.

## Supplementary Materials

**Figure 8.**
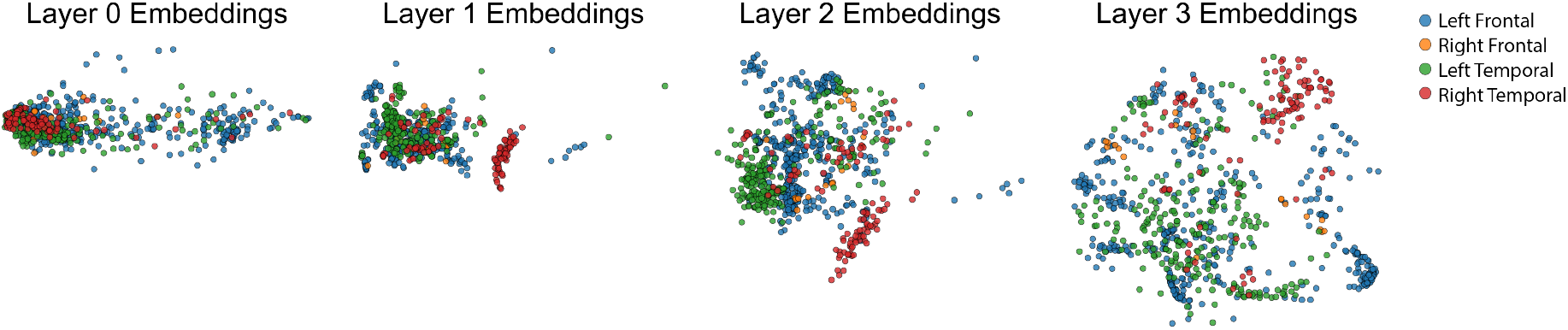
Projection of 2-dimensional PCA after each architectural block; each point represents a seizure. The embedding space visualization shows the distributions after processing through the 1D CNN tokenizer, the spatial encoder block, the temporal encoder block, and finally the SwAV projection head.

**Figure 9.**
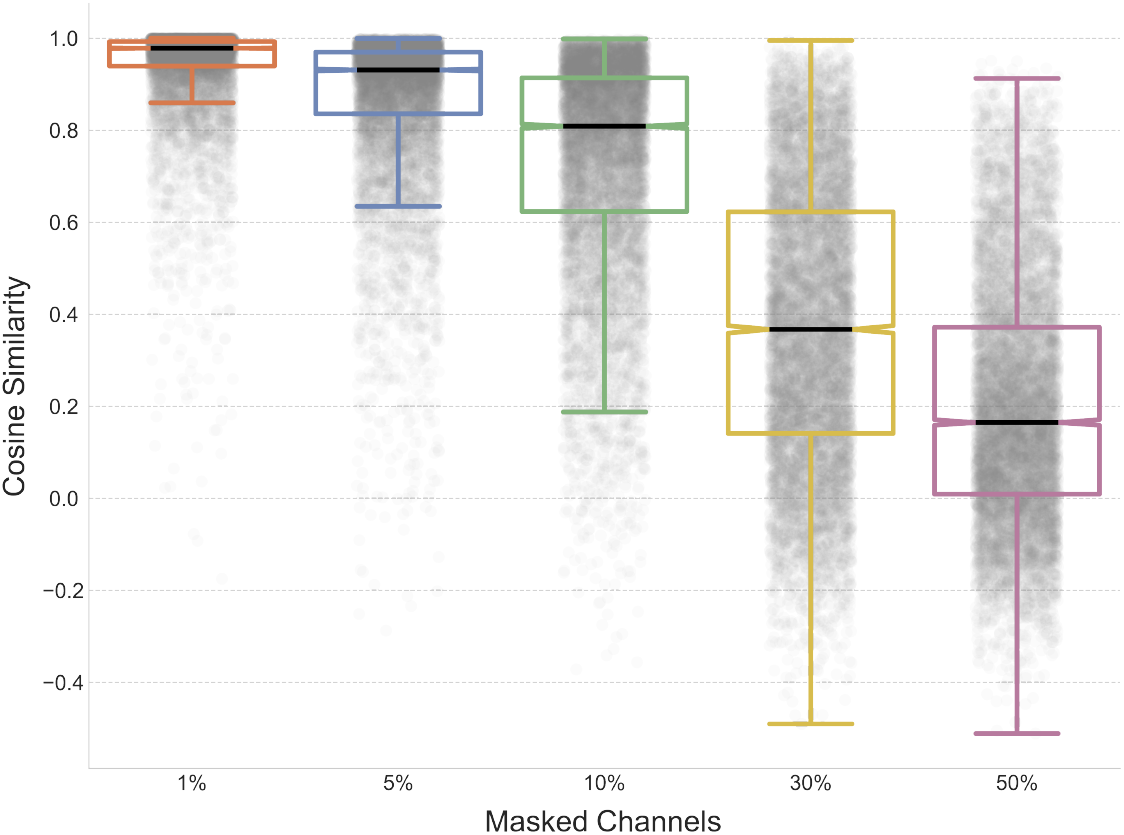
Cosine Similarity between Full Seizure Data and Masked Channels. Each point represents a cosine similarity score. We randomly mask a certain percentage of recording channels, pass the masked data through the trained model, and compute the cosine similarity between embeddings from the original data and those from masked channels. Each seizure was bootstrapped 10 times.

## Notes

### Competing Interest Statement

The authors have declared no competing interest.

### Author Declarations

Institutional Review Board of University of Pennsylvania gave ethical approval for this work

